# Domain-Specific Effects of GABA-Modulating Pharmacotherapies in Autism Spectrum Disorder: A Systematic Review and Meta-Analysis of Randomized Controlled Trials

**DOI:** 10.64898/2026.08.23.26361086

**Authors:** Sanjana Palakodeti, Kirti Kumar Hinduja, Garima Misra, R Subhiksha, Abhigna Pabbaraju, Bagath Srinivasan Balaji, Tejesvi Parmar

## Abstract

**Background:** Altered γ-aminobutyric acid (GABA) neurotransmission is a proposed mechanism underlying autism spectrum disorder (ASD), prompting evaluation of several GABA-modulating pharmacotherapies. However, it remains unclear whether these interventions improve ASD broadly or preferentially affect specific symptom domains.

**Methods:** We conducted a systematic review and random-effects meta-analysis of randomized controlled trials evaluating GABA-modulating pharmacotherapies in individuals with ASD. PubMed/MEDLINE, Embase, Scopus, and CENTRAL were searched from inception to April 1, 2026. Outcomes were prespecified as global autism severity, social communication, functional communication, restricted and repetitive behaviours (RRBs), adaptive behaviour, and irritability. Risk of bias was assessed using the Cochrane Risk of Bias 2 tool, and certainty of evidence was evaluated using GRADE.

**Results:** Thirteen randomized controlled trials evaluating three GABA-modulating interventions (bumetanide, arbaclofen, and valproate) were included. GABA-modulating therapies were associated with statistically significant improvements in global autism severity (Hedges’ g = −0.25, 95% CI −0.46 to −0.03; *p* = 0.028) and adaptive behaviour (Hedges’ g = −0.09, 95% CI −0.15 to −0.02; *p* = 0.023). No significant pooled effects were observed for social communication (Hedges’ g = −0.26, *p* = 0.077), functional communication (Hedges’ g = −0.01, *p* = 0.869), RRBs (Hedges’ g = −0.21, *p* = 0.126), or irritability (Hedges’ g = −0.07, *p* = 0.543). After Holm–Bonferroni step down procedure, neither global autism severity nor adaptive behaviour remained statistically significant (Holm-adjusted p = .140 and .138, respectively). Adverse events were predominantly gastrointestinal, neurological, metabolic, and appetite-related. Overall risk of bias was variable, and the certainty of evidence ranged from very low to moderate.

**Conclusions:** GABA-modulating pharmacotherapies did not demonstrate a robust, multiplicity-corrected benefit in any of the six prespecified ASD symptom domains. Nominal, unadjusted improvements in global autism severity and adaptive behaviour did not withstand correction for multiple comparisons and should be regarded as hypothesis-generating rather than confirmatory. Larger, adequately powered randomized trials using standardized domain-specific outcome measures are needed to determine whether individual GABA-modulating agents provide clinically meaningful benefit.

## Introduction

Autism Spectrum Disorder (ASD) is a heterogenous neurodevelopmental disorder that typically manifests in early childhood. It is characterized by impairments in social communication, repetitive and stereotyped behaviors, restricted interests, and sensory perception abnormalities (Wang and Sun, 2025). Approximately 1 in 31 (3.2%) children aged 8 years had ASD as per the estimates from CDC’s ADDM Network, which also suggested that it was 3 times more common among boys than among girls (Shaw, 2025). ASD frequently co-occurs with other neurodevelopmental and medical conditions, including global developmental delay, Attention Deficit Hyperactivity Disorder (ADHD), language disorders, epilepsy, and genetic syndromes, which contribute to substantial heterogeneity in clinical presentation and functional impairment across affected children (Burns et al., 2023). This heterogeneity is not incidental; it means that children carrying the same diagnostic label can present with entirely different dominant impairments: one child’s primary burden may be repetitive, restrictive behavior, while another’s is a near-absence of functional language, and a third’s is severe irritability that precludes any other intervention. Consequently, a treatment that produces little change in a global severity score may still have meaningful clinical value if it preferentially improves specific symptom domains.

An imbalance between excitatory (glutamatergic) and inhibitory (GABAergic) neurotransmission – the excitation/inhibition (E/I) imbalance hypothesis is one of the influential pathophysiological hypotheses in ASD. It is supported by genetic evidence (variants in GABA receptor subunit genes GABRB3, GABRA5, GABRG3, within 15q11–q13) and by developmental evidence (delayed maturation of the NKCC1/KCC2 chloride cotransporters that normally convert GABA’s action from excitatory to inhibitory in early life) (Zhao et al., 2022; Zhang et al., 2020). Rather than identifying a single therapeutic target, this hypothesis identifies a shared neurobiological mechanism that can be modulated through multiple pharmacological approaches.

Importantly, the therapeutic effects of GABAergic modulation in ASD are not expected to be unidirectional, and this bidirectionality has direct implications on how any “class-level” evidence on GABA-modulating drugs should be interpreted. While GABA receptor agonists and positive allosteric modulators may enhance inhibitory signalling, some individuals with ASD may experience paradoxical worsening of anxiety or aggression with agents such as benzodiazepines (Burns et al., 2023). This may reflect altered chloride homeostasis and a shift in GABAergic signalling from inhibitory toward excitatory. Restoring the inhibitory action of GABA through NKCC1 inhibition with bumetanide represents an alternative approach to targeting the same underlying E/I imbalance (Burns et al., 2023). Thus, both enhancement and normalization of GABAergic signalling may have therapeutic potential in ASD, but they may also act on different patients, in different directions, and to different degrees. This provides both the rationale for evaluating diverse GABA-modulating pharmacotherapies as a broader mechanistically related treatment strategy, and a reason to expect that pooling agents with different mechanisms of action could dilute or obscure true effects that exist for any single agent (Cellot and Cherubini, 2014).

Accordingly, clinical trials have evaluated diverse GABA-modulating interventions, including GABA-B receptor agonists (arbaclofen, baclofen), restoration of chloride-dependent inhibitory signalling (bumetanide), and broader GABAergic modulators (valproate, topiramate, cenobamate, benzodiazepines, and barbiturates). Studies investigating GABA-modulating therapies in ASD suggest that Bumetanide may improve core ASD symptoms, although adverse effects such as hypokalemia, dehydration, and diuresis have been frequently reported. Arbaclofen has shown promise in improving social functioning, whereas evidence for Valproate remains inconsistent, particularly for irritability and aggression. Overall, despite encouraging therapeutic signals, concerns regarding tolerability and variability in clinical outcomes continue to limit the broader application of GABA-targeting therapies in ASD (Lemonnier et al., 2012; Lemonnier et al., 2017; Wang and Sun, 2025; Frye, 2014; Valle et al., 2025)

Prior reviews of GABA-modulating agents in ASD have been limited in scope or resolution. Brondino et al. (2015) conducted a systematic review of GABAergic drugs (valproate, acamprosate, arbaclofen) but reported findings qualitatively, without pooling effect sizes, and concluded that evidence was insufficient to guide clinical use (Brondino et al., 2015). Subsequent quantitative work has focused on single agents in isolation, such as bumetanide, thereby estimating the efficacy of single pharmacological agents rather than the broader therapeutic strategy of GABA modulation (Wang and Sun, 2025). Conversely, broader network meta-analyses have compared numerous pharmacological interventions for ASD and stratified treatment effects according to symptom domains, but they evaluate GABA-modulating agents as separate interventions within a network of mechanistically unrelated therapies rather than as a mechanistically related treatment strategy (Siafis et al., 2022).

Consequently, two questions remain unanswered. First, whether pharmacological modulation of GABAergic signalling produces consistent improvements across all ASD symptom domains or preferentially benefits selected domains of impairment. Second, given mechanistic heterogeneity, whether any overall effect reflects a shared benefit of GABA modulation or is mainly driven by one or a few specific agents. To address this knowledge gap, we performed a systematic review and meta-analysis of randomized controlled trials evaluating GABA-modulating pharmacotherapies in ASD across six prespecified clinical domains: global autism severity, social communication, functional communication, restricted and repetitive behaviours, adaptive functioning, and irritability to provide a comprehensive evaluation of the therapeutic effects of GABA-modulating therapies across ASD symptom domains.

## Methodology

### 2.1 Study Design and Protocol Registration

This systematic review and meta-analysis were conducted in accordance with the Preferred Reporting Items for Systematic Reviews and Meta-Analyses (PRISMA) 2020 guidelines (Page et al., 2021) and the Cochrane Handbook for Systematic Reviews of Interventions (Higgins et al., 2024). The review protocol was prospectively registered with the International Prospective Register of Systematic Reviews (PROSPERO) (Registration number: CRD420261451049) (Palakodeti et al., 2026).

### 2.2 Eligibility Criteria

Studies were considered eligible if they: (1) enrolled children, adolescents, or adults with a diagnosis of autism spectrum disorder established using standardized diagnostic criteria or validated diagnostic instruments; (2) evaluated at least one GABA-modulating pharmacotherapy (arbaclofen, baclofen, bumetanide, riluzole, valproate, topiramate, cenobamate, benzodiazepines, or barbiturates); (3) included a placebo comparator; (4) reported quantitative data for at least one predefined outcome; and (5) employed a randomized controlled trial design. Studies were excluded if they: (1) were non-randomized studies, observational studies, case series, conference abstracts, reviews, editorials, or case reports; (2) lacked a placebo comparator; (3) enrolled participants without a formal diagnosis of autism spectrum disorder; or (4) did not report sufficient data for effect size calculation.

### 2.3 Search Strategy and Study Selection

A systematic literature search was conducted in PubMed/MEDLINE, Embase, Scopus, and the Cochrane Central Register of Controlled Trials (CENTRAL) from database inception until 1 April 2026. The search strategy included synonyms and variations of autism spectrum disorder together with eligible interventions, including arbaclofen, baclofen, bumetanide, riluzole, valproate, topiramate, cenobamate, benzodiazepines, and barbiturates. The complete search strategies for all databases are provided in Supplementary Table S1.

All retrieved records were imported into Rayyan systematic review software, and duplicate records were removed before screening. Two reviewers independently screened titles and abstracts for eligibility, followed by full-text review of potentially eligible articles. During full-text screening, additional duplicate reports were identified, including instances in which trial registrations or other reports corresponded to an already included full-text publication; these were treated as reports of the same study rather than independent studies. Disagreements were resolved through consensus, with consultation of a third reviewer where necessary. To enhance the completeness of the search, reference lists of included studies and relevant recent systematic reviews and meta-analyses were additionally screened for potentially eligible articles not captured through the electronic database searches. A targeted supplementary search for valproate was also conducted based on studies identified from a recent systematic review and meta-analysis of antiepileptic drugs in ASD, with additional searches covering the period from 2024 to April 2026. This approach was used to identify potentially eligible recent studies while limiting the retrieval of a large volume of irrelevant records associated with broader valproate searches. The study selection process was documented using a PRISMA flow diagram.

### 2.4 Data Extraction

Two reviewers independently extracted data using a Standardized data extraction form. Extracted variables included study characteristics, participant demographics, intervention details, treatment duration, comparator characteristics, and outcome data. For continuous outcomes, means and standard deviations at baseline and endpoint were preferentially extracted. For studies reporting multiple intervention arms with a shared placebo group, intervention arms were combined before comparison with the placebo group to avoid double counting of control participants. For studies reporting only participant-level outcome data, summary statistics were calculated manually in Microsoft Excel before inclusion in the quantitative synthesis. Safety data, including reported adverse events, were also extracted from all included studies. To provide an overview of the safety profile of GABA-modulating interventions, a descriptive summary table of adverse events reported in three or more studies was prepared.

### 2.5 Outcomes

Outcomes were prespecified and categorized into five primary domains reflecting core ASD symptom dimensions and functional outcomes: **global ASD symptom severity, social functioning/social communication, functional communication, restricted and repetitive behaviours (RRBs), and adaptive functioning**. Secondary outcomes included **irritability symptoms**. Conceptually similar instruments assessing the same construct were pooled within their respective domains using standardized mean differences (Hedges’ g), allowing quantitative pooling across studies employing different outcome measures. The allocation of individual outcome measures to each domain and its rationale are provided in Supplementary Tables S2 & S3.

When multiple outcome measures from the same study contributed to a single outcome domain, only one measure was included in the primary analysis to maintain independence of observations. Selection was based on a prespecified hierarchical framework that prioritized: (1) construct-specific instruments; (2) autism-specific measures over general behavioural or adaptive measures; (3) symptom severity scales over diagnostic algorithm subscales; (4) clinician-rated measures over caregiver-rated measures when construct coverage was comparable; and (5) the most frequently used instrument across included studies when multiple measures remained equivalent. The complete hierarchy and study-specific selection decisions are presented in Supplementary Tables S4 and S5.

### 2.6 Statistical Analysis

Continuous outcomes were synthesized using standardized mean differences (Hedges’ g) with 95% confidence intervals (CIs), while dichotomous outcomes were summarized using risk ratios (RRs) with 95% CIs. Random-effects meta-analyses was conducted using the Hartung–Knapp adjustment for inference. For outcome measures in which higher scores indicated better functioning, effect sizes were reversed before pooling to ensure a consistent direction of effect, with negative values favoring the intervention. Statistical heterogeneity was assessed using Cochran’s Q test and quantified using the I² statistic, with values of 25%, 50%, and 75% representing low, moderate, and high heterogeneity, respectively. Sensitivity analyses were performed where appropriate, and publication bias was assessed by visual inspection of funnel plots when sufficient studies were available. All analyses were conducted using JASP version 0.98.1. A p-value < 0.05 was considered statistically significant for all analyses.

Because six outcome domains were evaluated, a post hoc sensitivity analysis was conducted to assess the potential impact of multiple comparisons on the interpretation of the meta-analytic findings. The six outcome-level p-values from the primary random-effects meta-analyses were treated as a single family of hypotheses and adjusted using the Holm–Bonferroni step-down procedure. The unadjusted p-values were ranked from smallest to largest, and Holm-adjusted p-values were calculated sequentially according to the rank of each test. Statistical significance after adjustment was defined as a Holm-adjusted p-value < .05. This post hoc adjustment was applied to the p-values only and did not alter the pooled effect estimates or their corresponding 95% confidence intervals. The Holm-adjusted p values are detailed in Supplementary Table S6.

### 2.7 Risk of Bias Assessment

Methodological quality of included studies was independently assessed by two reviewers using the Cochrane Risk of Bias 2 (RoB 2) tool for randomized controlled trials (Sterne et al., 2019). Disagreements were resolved through discussion and consensus. The assessment evaluated potential sources of bias arising from the randomization process, deviations from intended interventions, missing outcome data, outcome measurement, and selection of the reported results. Each domain was judged as having low risk of bias, some concerns, or high risk of bias according to the RoB 2 guidance. The results were summarized using graphical representations, including a summary plot to illustrate the distribution of bias across studies

### 2.8 Certainty of evidence (GRADE)

The certainty of evidence for each prespecified outcome was assessed using the Grading of Recommendations Assessment, Development and Evaluation (GRADE) approach and the GRADEpro GDT Tool (Schünemann et al., 2013; McMaster University and Evidence Prime, 2023). GRADE certainty was evaluated across five domains: risk of bias, inconsistency, indirectness, imprecision, and other considerations, including potential publication bias. The assessment was performed for the pooled estimates of global autism severity, social communication, functional communication, restricted and repetitive behaviours (RRBs), adaptive behaviour, and irritability. The overall certainty for each outcome was classified as high, moderate, low, or very low.

## Results

### 3.1 Study selection and characteristics

The study selection process is presented in Figure 1. The systematic search identified 865 records from 4 databases. After screening, ten studies met the eligibility criteria and were included in the updated systematic review and meta-analysis. Together with three studies from a broader systematic review on anti-epileptics for ASD, a total of thirteen studies were included in the final analysis. The included studies evaluated three GABA-modulating interventions: bumetanide (n = 8) (Dai et al., 2021; Lemonnier et al., 2012; Lemonnier et al., 2017; Sprengers et al., 2021; Du et al., 2015; Zhang et al., 2020; Fuentes et al., 2023) arbaclofen (n = 2) (Parellada et al., 2026; Veenstra-VanderWeele et al., 2016), and valproate (n = 3) (Hollander et al., 2010; Hollander et al., 2005, Hellings et al., 2005). All the studies included were randomised controlled trials. A summary of the characteristics of the included studies is presented in Table 1.

**Figure 1.**
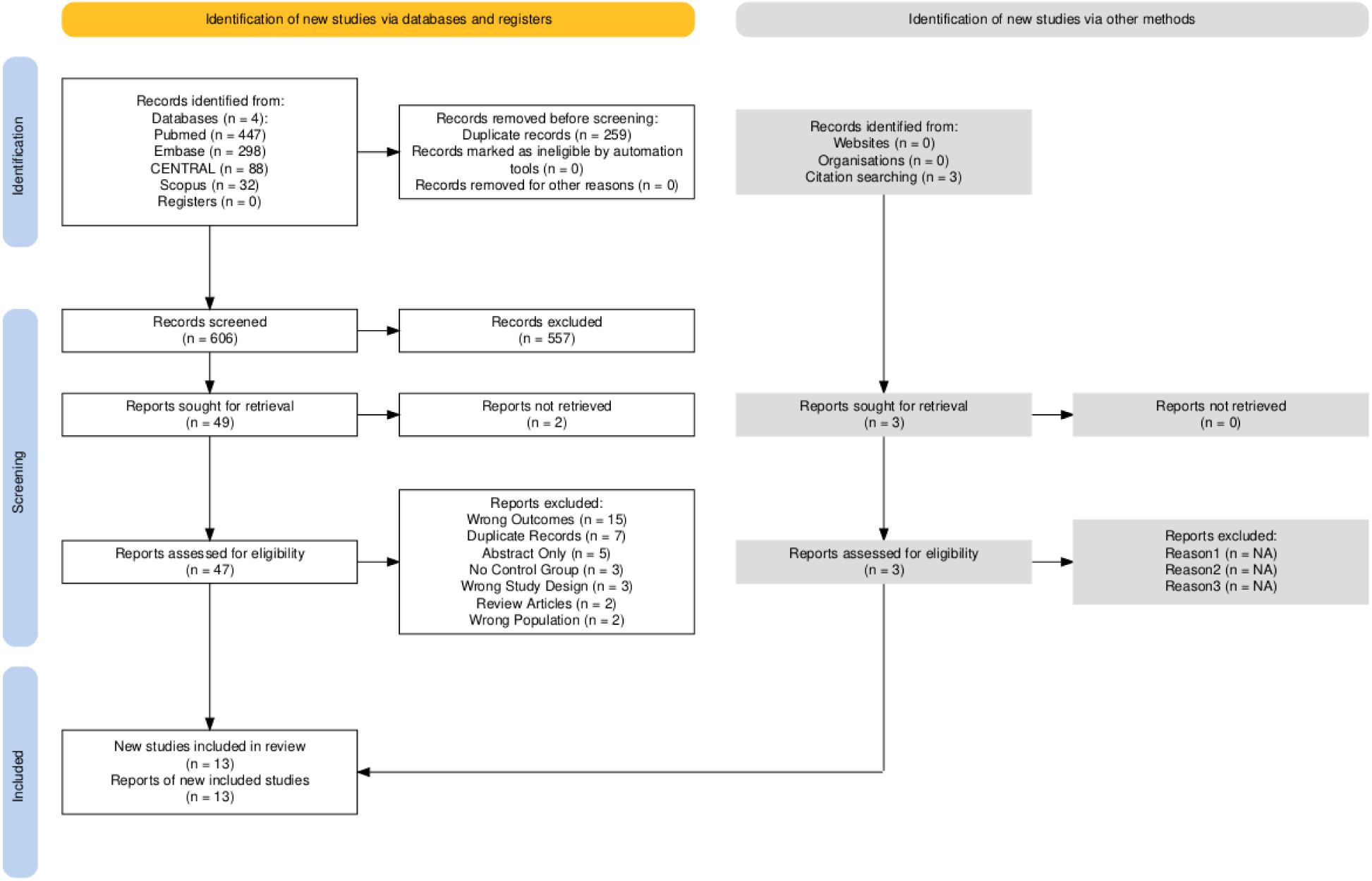
PRISMA 2020 flow diagram of study selection.

**Table 1:** Baseline study and patient characteristics.

| Study, Year | Study design | Country | Particip<br>ants,<br>I/C | Age<br>(Years),<br>Mean ±<br>SD | Sex,<br>M/F | GABA<br>Modulator | Dose | Treatment<br>Duration** |
| --- | --- | --- | --- | --- | --- | --- | --- | --- |
| Parellada et al.,<br>2026 | Phase II,<br>randomized,<br>double-blind,<br>placebo-controlled | Spain | 59/63 | 11.67 ±<br>3.20 | 102/20 | Arbaclofen | Age-based<br>titration (5–20 mg<br>TID) | 16 weeks |
| Vanderweele et al.,<br>2016 | Phase II,<br>randomized,<br>double-blind,<br>placebo-controlled<br>, multisite | USA | 76/74 | 11.6 ±<br>4.6 | 124/26 | Arbaclofen | 26.8 mg/day<br>(children); 41.7<br>mg/day (adults) | 12 weeks |
| Fuentes A et al.,<br>2023 | Phase III,<br>randomized,<br>double-blind,<br>placebo-controlled | Multinational | 107/104 | 10.45 ±<br>2.94 | 174/37 | Bumetanide | 0.02 mg/kg BID<br>(<25 kg); 0.5 mg<br>BID (≥25 kg) | 6 months |
| Fuentes B et al.,<br>2023 | Phase III,<br>randomized,<br>double-blind,<br>placebo-controlled | Multinational | 107/104 | 4.50 ±<br>1.15 | 176/35 | Bumetanide | 0.02 mg/kg BID<br>(<25 kg); 0.5 mg<br>BID (≥25 kg) | 6 months |
| Dai et al., 2021 | Randomized,<br>double-blind,<br>placebo-controlled,<br>parallel-group | China | 59/60 | 4.12 | 100/19 | Bumetanide | 0.5 mg BID | 3 months |
| Sprengers et al., 2021 | Phase II,<br>randomized,<br>double-blind,<br>placebo-controlled | Netherlands | 47/45 | 10.5 ± 2.4 | 63/29 | Bumetanide | 0.5–1.0 mg BID | 13 weeks |
| Zhang et al., 2020 | Randomized,<br>open-label,<br>controlled | China | 42/41 | 4.09 ± 0.98 | 65/18 | Bumetanide | 0.5 mg BID | 3 months |
| Lemonnier et al., 2017 | Phase II,<br>randomized,<br>double-blind,<br>placebo-controlled<br>, dose-ranging | France | 65*/23 | 8.26 ± 4.44 | 78/10 | Bumetanide | 0.5, 1.0, or 2.0 mg<br>BID | 3 months |
| Du et al., 2015 | Pilot study | China | 28/32<sup>^</sup> | 4.55 ± 1.78 | 51/9 | Bumetanide | 0.5 mg BID | 3 months |
| Lemonnier et al., 2012 | Randomized,<br>placebo-controlled | France | 30/30 | 6.49 ± 1.89 | NR | Bumetanide | 1 mg/day (0.5 mg<br>BID) | 3 months |
| Hollander et al.,<br>2010 | Randomized,<br>double-blind,<br>placebo-controlled | USA | 16/11 | 9.46 ±<br>2.65 | 23/4 | Divalproex<br>sodium | Weight-adjusted<br>(target serum ≥50<br>µg/mL) | 12 weeks |
| Hellings et al.,<br>2005 | Double-blind,<br>placebo-controlled | USA | 16/14 | 11.2 ±<br>4.2 | 22/8 | Divalproex<br>sodium | Titrated to 20<br>mg/kg/day | 8 weeks |
| Hollander et al.,<br>2005 | Randomized,<br>double-blind,<br>placebo-controlled | USA | 9/4 | 9.5 (SD<br>NR) | NR | Divalproex<br>sodium | 125 mg/day,<br>titrated by 125 mg<br>every 4 days | 8 weeks |

I/C - Number in intervention group/ number in control group
M/F - Number of males/ Number of females
BID - Twice a day
TID - Thrice a day
NR - Not reported
\*- Three bumetanide intervention arms (0.5 mg BID, n = 20; 1.0 mg BID, n = 23; 2.0 mg BID, n = 22) were pooled as a single intervention group for analysis.
\*\* - Treatment duration refers to the duration of the randomized intervention phase included in the meta-analysis. Open-label extensions, taper periods, and washout periods are not reflected unless they formed part of the randomized study design.
^ - 5 participants subsequently dropped out, and outcome analyses were reported for 55 participants (26 intervention, 29 control).

### 3.2 Outcomes

The quantitative synthesis evaluated six outcome domains. Forest plots for Global Autism Severity, Social Communication, and Functional Communication are presented in Figure 2, whereas forest plots for Restricted and Repetitive Behaviours (RRBs), Adaptive Behaviour, and Irritability are presented in Figure 3.

**Figure 2.**
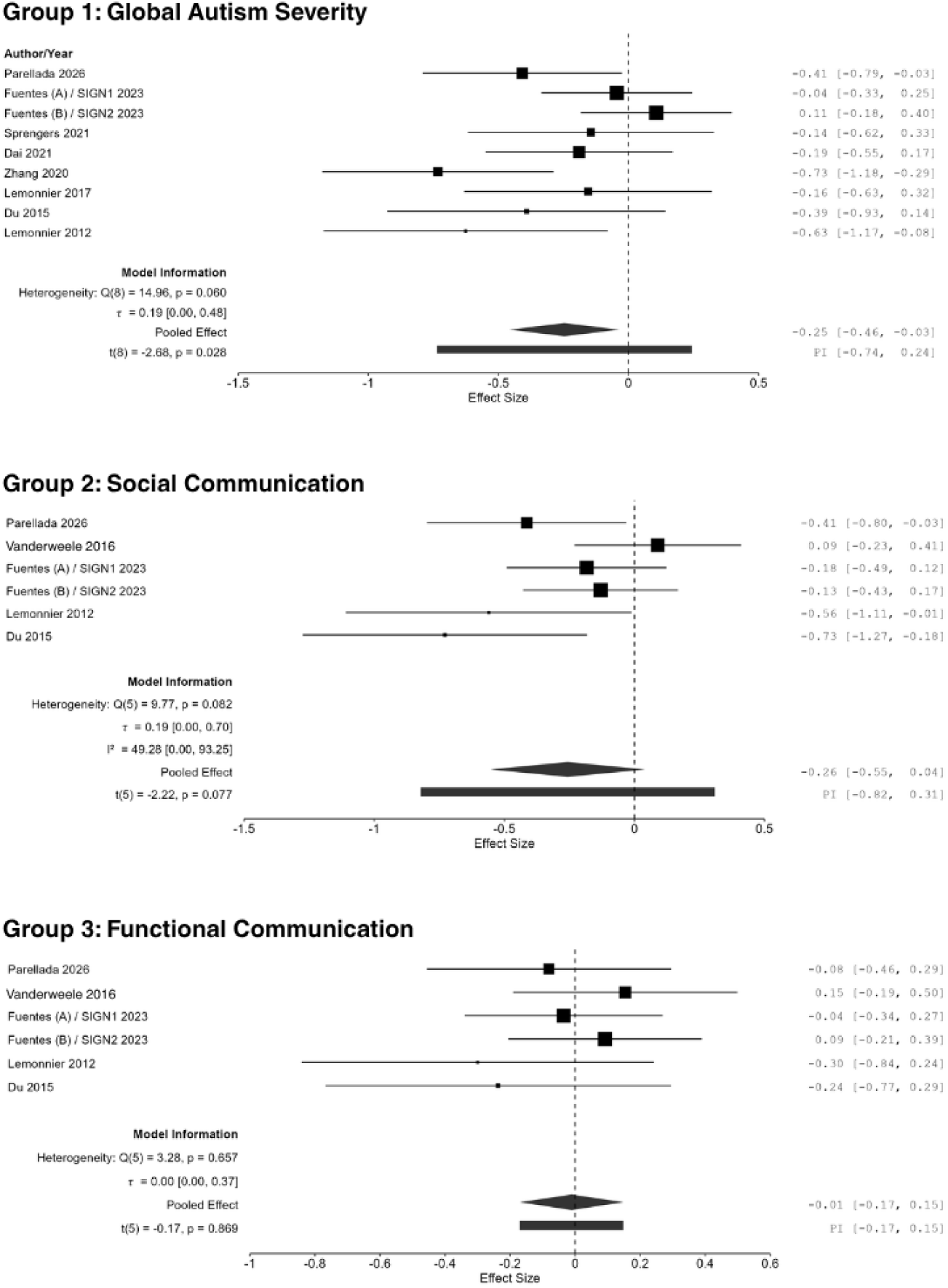
Forest plots of the effects of GABA-modulating interventions on global autism severity, social communication, and functional communication.

**Figure 3.**
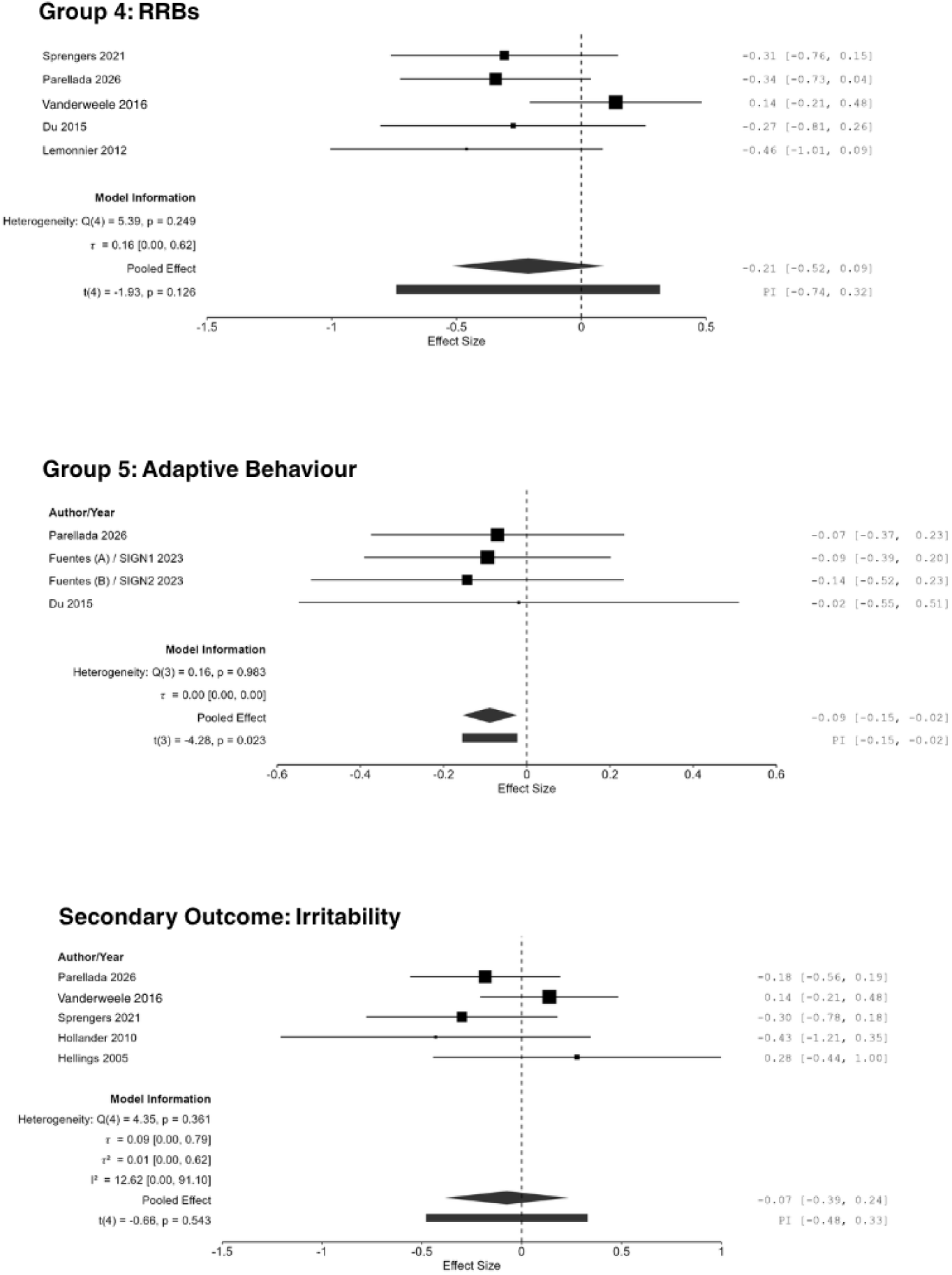
Forest plots of the effects of GABA-modulating interventions on restricted and repetitive behaviours, adaptive behaviour, and irritability.

#### 3.2.1 Global autism severity

Nine study estimates were included in the global autism severity analysis. The pooled effect favored the intervention and reached nominal statistical significance (Hedges’ g = −0.25, 95% CI −0.46 to −0.03; t(8) = −2.68, p = .028). Heterogeneity was moderate but not statistically significant (Q(8) = 14.96, p = .060; I² = 47.38%). A sensitivity analysis excluding Zhang et al. (2020), the only open-label trial, attenuated the pooled effect and rendered it statistically nonsignificant (Hedges’ g = −0.18, 95% CI −0.36 to 0.01; t(7) = −2.19, p = .065) (Supplementary Figure S1). Heterogeneity was reduced (Q(7) = 9.01, p = .252; I² = 26.1%). Overall, the pooled estimate favored the intervention, although the nominally significant finding was sensitive to exclusion of the open-label trial.

Visual inspection of the funnel plot revealed asymmetry. Formal tests of funnel plot asymmetry yielded statistically significant results (meta-regression test: z = −2.78, p = .005; weighted regression/Egger-type test: t = −2.728, p = .029), indicating possible small-study effects. These findings should be interpreted cautiously given the limited number of available study estimates (n = 9), as formal asymmetry tests have limited power and may be influenced by factors other than publication bias. (Supplementary Figure S2)

#### 3.2.2 Social Communication

Six studies contributed to the social communication domain. The pooled effect was not statistically significant (Hedges’ g = -0.26, 95% CI -0.55 to 0.04; t(5)=-2.22, p=.077). Heterogeneity was moderate (Q(5)=9.77, p=.082; I^2^=49.28%). The available evidence did not demonstrate a meaningful effect on social communication.

#### 3.2.3 Functional communication

Six studies contributed to the functional communication domain. The pooled effect was not statistically significant (Hedges’ g = -0.01, 95% CI -0.17 to 0.15; t(5)=-0.17, p=.869). Heterogeneity was low to moderate (Q(5)=3.28, p=.657; I^2^=25.94%). Thus, the available evidence did not demonstrate a meaningful effect on functional communication.

#### 3.2.4 Restricted and repetitive behaviours

Five studies were included in the meta-analysis of restricted and repetitive behaviours. Lemonnier et al. (2017) was excluded because the study reported change scores but did not provide baseline and endpoint scores, and therefore an endpoint estimate could not be calculated. The pooled effect favored the intervention but was not statistically significant (Hedges’ g = -0.21, 95% CI -0.51 to 0.09; t(4)=-1.93, p=.126). Heterogeneity was low to moderate (Q(4)=5.39, p=.249; I^2^ = 32.54%). These results suggest a possible reduction in restricted and repetitive behaviours, but the evidence was not definitive. One additional study (Hollander et al., 2005) could not be included in the quantitative synthesis because C-YBOCS outcome data were presented graphically without sufficient numerical data for effect-size calculation. However, the study reported a statistically significant between-group improvement in repetitive behaviours at 8 weeks (p = .037; Cohen’s d = 1.616).

#### 3.2.5 Adaptive behaviour

Four studies contributed to the adaptive behaviour analysis. The pooled effect was statistically significant and favored the intervention (Hedges’ g = -0.09, 95% CI -0.15 to -0.02; t(3)=-4.28, p=.023). Heterogeneity was negligible (Q(3)=0.16, p=.983; I^2^=0.00%) . This indicates a small but consistent benefit for adaptive functioning.

#### 3.2.6 Irritability

Five studies contributed data to analyse irritability scores using the ABC-Irritability score. The random-effects model showed no significant pooled effect (Hedges’ g = -0.07, 95% CI -0.39 to 0.24; t(4)=-0.66, p=.543), and heterogeneity was low (Q(4)=4.35, p=.361; I^2^=12.62%). These findings suggest that the interventions did not demonstrate a clear improvement in irritability.

#### 3.2.7 Safety outcomes

Adverse events reported across the included studies were primarily gastrointestinal, neurological, metabolic, and appetite-related. Frequently reported events included abdominal pain, diarrhea, nausea, vomiting, headache, decreased appetite, hypokalemia, and polyuria. Several adverse events were reported across more than one GABA-modulating intervention. A descriptive summary of the reported adverse events is presented in Table 2.

**Table 2.** Summary of Frequently Reported Adverse Events.

| Adverse Event | Studies Reporting, n | Intervention Events, n | Intervention Participants, n | Control Events, n | Control Participants, n | GABA-Modulating Agents Reporting the Event |
| --- | --- | --- | --- | --- | --- | --- |
| Diarrhea | 6 | 23 | 300 | 22 | 256 | Bumetanide, Valproate, Arbaclofen |
| Polyuria | 6 | 93 | 259 | 9 | 153 | Bumetanide, Valproate |
| Vomiting | 6 | 43 | 300 | 25 | 256 | Bumetanide, Valproate, Arbaclofen |
| Headache | 5 | 43 | 213 | 48 | 174 | Bumetanide, Valproate, Arbaclofen |
| Hypokalemia | 5 | 104 | 396 | 6 | 303 | Bumetanide |
| Decreased appetite | 5 | 34 | 300 | 13 | 192 | Bumetanide, Arbaclofen |
| Abdominal pain | 4 | 31 | 185 | 21 | 120 | Bumetanide, Valproate, Arbaclofen |
| Nausea | 4 | 22 | 180 | 18 | 180 | Bumetanide, Valproate, Arbaclofen |
| Drowsiness | 3 | 13 | 108 | 3 | 61 | Valproate, Arbaclofen |
| Fatigue/Asthenia | 3 | 14 | 152 | 5 | 65 | Bumetanide |
| Hyperuricemia | 3 | 8 | 165 | 1 | 82 | Bumetanide |
| Weight gain | 3 | 12 | 91 | 6 | 88 | Valproate, Arbaclofen |
| Weight loss | 3 | 13 | 179 | 3 | 139 | Bumetanide, Arbaclofen |

n, number of events or participants
**Notes:** Event counts represent reported adverse events and may exceed the number of affected participants. Only adverse events reported in $\geq 3$ included studies are presented.

### 3.3 Adjustment for multiple comparisons

As a post hoc sensitivity analysis, p-values were adjusted for multiple comparisons across the six outcome domains using the Holm–Bonferroni procedure. The nominally significant findings for global autism severity (unadjusted p = .028; Holm-adjusted p = .140) and adaptive behaviour (unadjusted p = .023; Holm-adjusted p = .138) did not remain statistically significant after adjustment. The remaining outcomes were also nonsignificant after adjustment (social communication, p = .308; restricted and repetitive behaviours, p = .378; irritability, p = 1.000; functional communication, p = 1.000). Thus, none of the six outcome domains remained statistically significant after controlling for multiplicity. The Holm-adjusted p values are available in Supplementary Table S6.

### 3.4 Risk of Bias Assessment

The risk of bias of the included randomized controlled trials was assessed using the Cochrane Risk of Bias 2 (RoB 2) tool. Overall, methodological quality was variable, with only two studies judged to be at low overall risk of bias, four studies rated as having some concerns, and seven studies judged to be at high risk of bias. Most studies demonstrated a low risk of bias in the domains of randomization (D1), deviations from intended interventions (D2), and missing outcome data (D3), indicating that the included trials generally employed appropriate randomization procedures, maintained intervention fidelity, and had limited attrition. In contrast, greater concerns were identified in the domains of measurement of the outcome (D4) and particularly selection of the reported result (D5), where several studies were judged to have either some concerns or a high risk of bias. These findings suggest that selective outcome reporting and, to a lesser extent, outcome assessment represented the principal methodological limitations across the included evidence. Although the majority of studies were at low risk of bias in individual methodological domains, the accumulation of concerns across multiple domains resulted in nearly 85% of studies being judged as having either some concerns or a high overall risk of bias. Consequently, the pooled estimates should be interpreted with appropriate caution, as the overall certainty of the evidence may be influenced by methodological limitations in the primary studies. The distribution of risk-of-bias judgments across studies and domains is presented in Figure 4.

**Figure 4.**
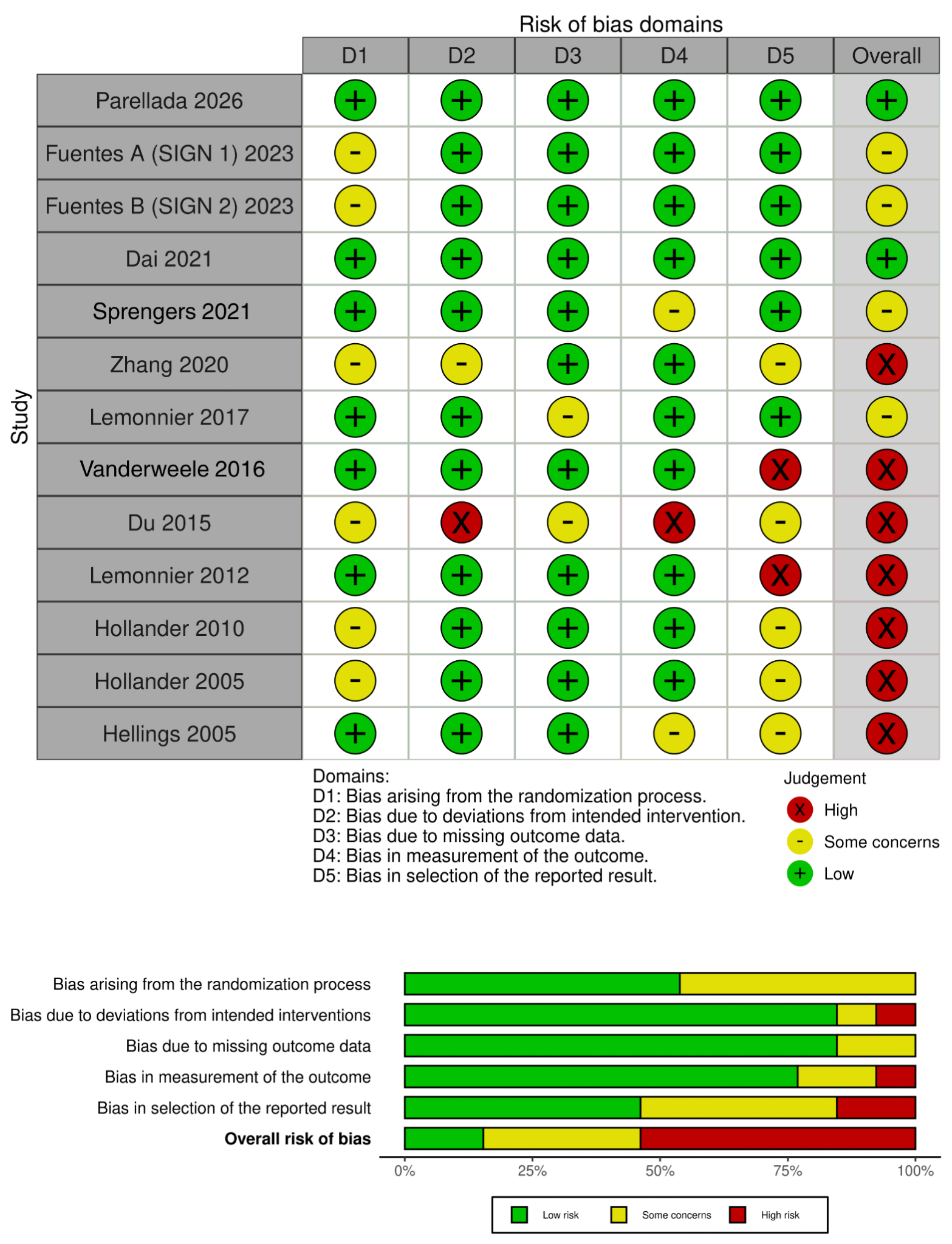
Risk of bias assessment of included studies using the Cochrane Risk of Bias 2 (RoB 2) tool. (A) Study-level risk of bias across the five RoB 2 domains and overall judgment. (B) Summary of the proportion of studies rated as low risk, some concerns, or high risk for each domain and the overall risk of bias.

### 3.5 Certainty of evidence (GRADE)

The certainty of evidence was moderate for global autism severity and adaptive behavior, low for functional communication, restricted and repetitive behaviors, and irritability, and very low for social communication. Downgrading was primarily related to concerns regarding risk of bias, with additional concerns regarding heterogeneity and imprecision for some outcomes. Detailed GRADE assessments are presented in Table 3.

**Table 3:**
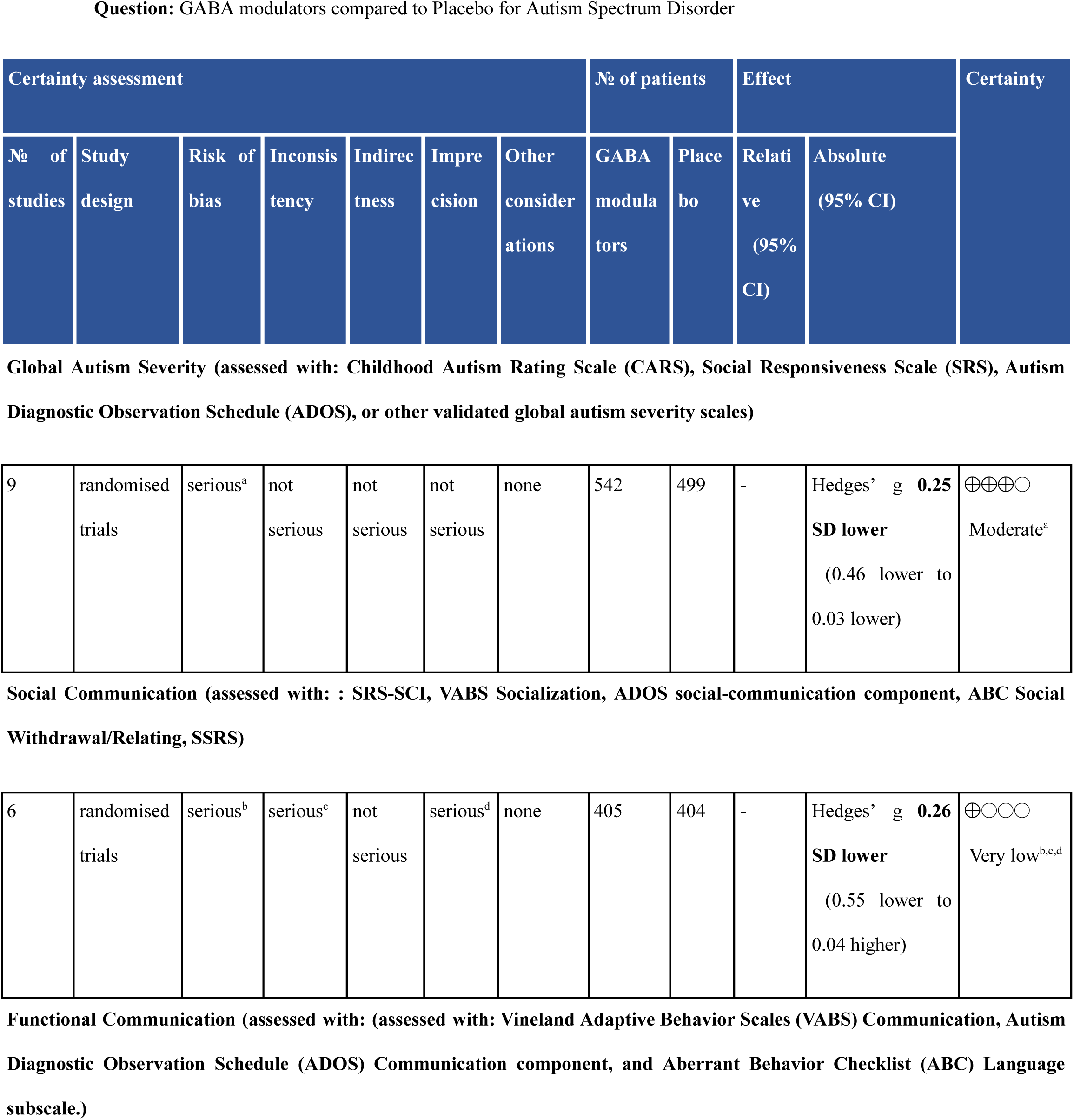
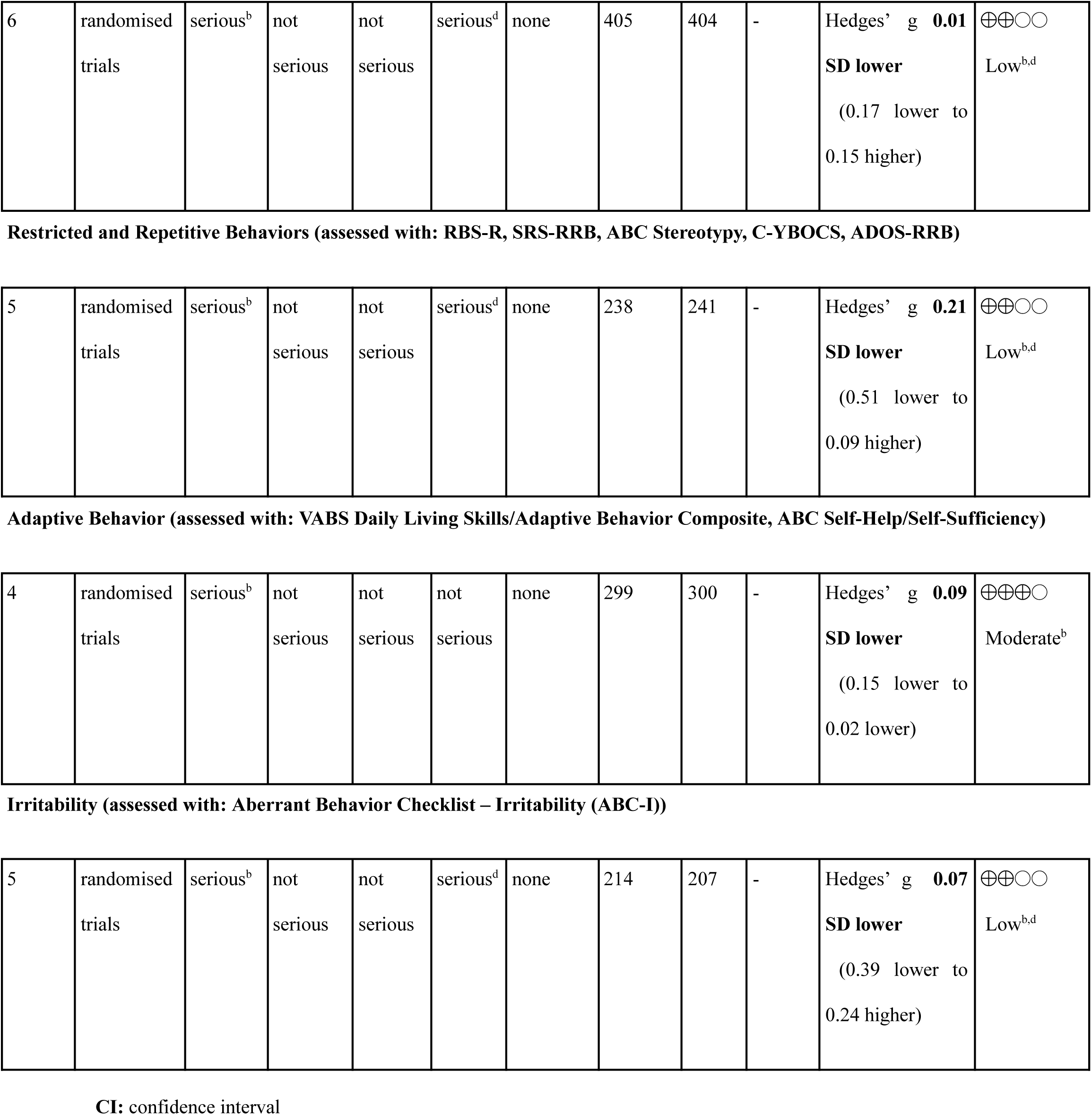
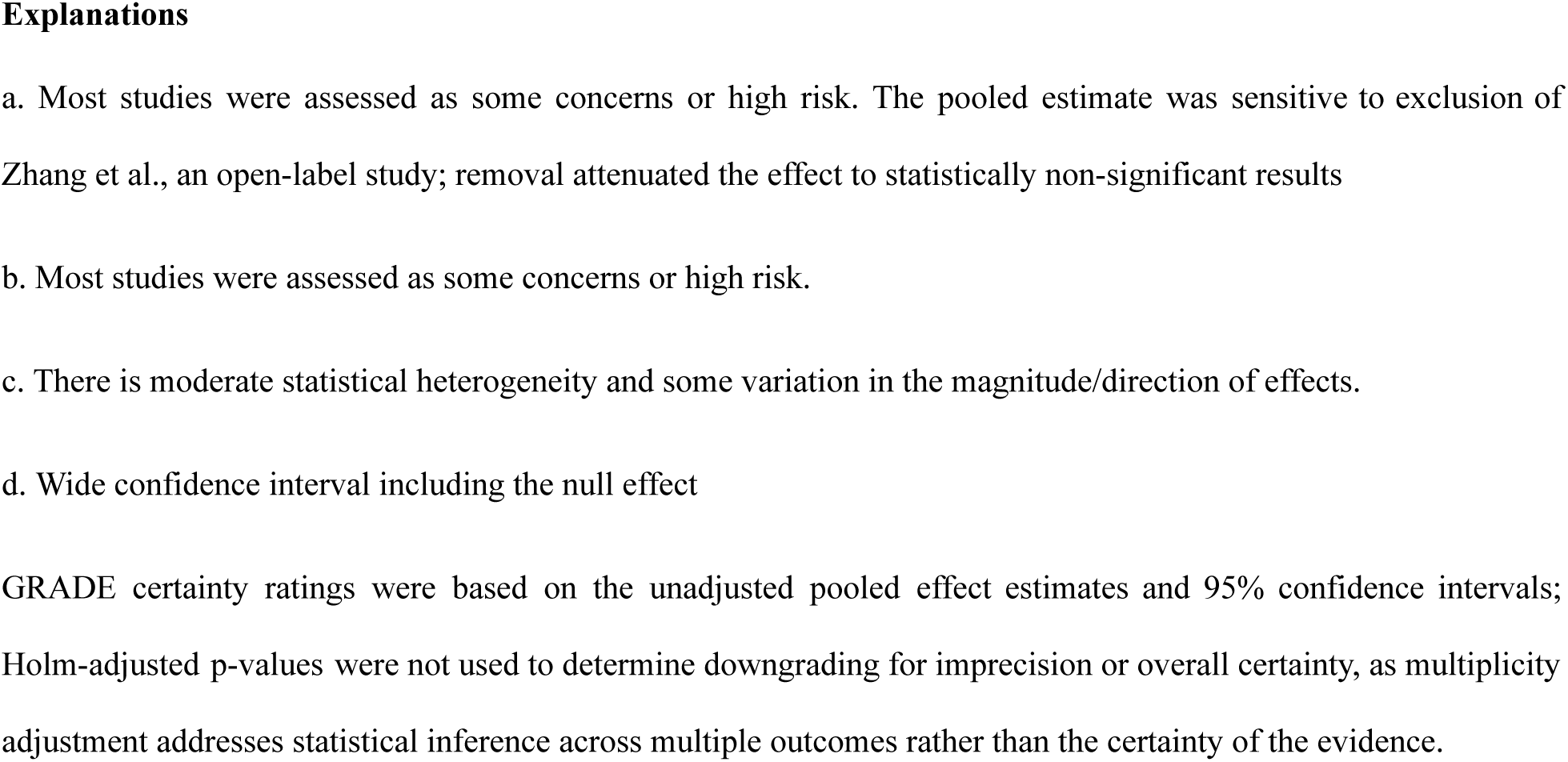
GRADE assessment. **Question:** GABA modulators compared to Placebo for Autism Spectrum Disorder

## Discussion

This systematic review and meta-analysis evaluated the efficacy and safety of GABA-modulating agents for autism spectrum disorder (ASD) across six prespecified domains. After accounting for multiple comparisons across these domains using the Holm–Bonferroni procedure, no domain reached statistical significance. Two domains — global autism severity and adaptive behaviour — showed nominally significant unadjusted effects (Hedges’ g = −0.25, p = .028, and g = −0.09, p = .023, respectively), but neither survived correction (Holm-adjusted p = .140 and .138). No significant pooled effects, adjusted or unadjusted, were observed for social communication, functional communication, restricted and repetitive behaviours (RRBs), or irritability. We regard the corrected result — that no domain shows a robust, multiplicity-adjusted signal — as the primary interpretive finding of this review, and we discuss the nominal, uncorrected findings below as hypothesis-generating rather than confirmatory.

### 4.1 The nominal signals for global severity and adaptive behaviour do not withstand scrutiny

Three independent factors argue against treating the nominal global-severity and adaptive-behaviour findings as reliable. First, neither remained statistically significant after Holm–Bonferroni correction for the six outcome domains tested, which is the appropriate correction given that all six domains were prespecified and tested within the same review. Second, the global-severity estimate was sensitive to the inclusion of a single open-label, unblinded trial (Zhang et al., 2020): excluding it attenuated the pooled effect and rendered it non-significant even before correction (Hedges’ g = −0.18, 95% CI −0.36 to 0.01; p = .065), which is consistent with an unblinding-related inflation of the pooled estimate rather than a genuine treatment effect. Third, formal tests of funnel-plot asymmetry for global severity were statistically significant (Egger-type test, p = .029), raising the possibility of small-study effects or selective reporting, a concern reinforced by the fact that bias in selection of the reported result (RoB 2 Domain 5) was the single most common risk-of-bias concern across the included trials. Taken together, these findings indicate that the apparent “selective” benefit for global severity is fragile and should not be presented as an established finding.

The adaptive-behaviour result is comparatively more internally consistent (I² = 0%, indicating no observed between-study heterogeneity) but rests on only four studies and an unadjusted p-value close to the multiple-comparisons-corrected threshold; with only four studies contributing, the review is underpowered to detect heterogeneity, so the absence of observed heterogeneity should not be over-interpreted as evidence of a uniform true effect. The estimated effect (Hedges’ g = −0.09) is also very small in magnitude, and its clinical, as opposed to statistical, significance remains uncertain.

### 4.2 Class-level pooling of mechanistically distinct agents limits interpretation

An important limitation of this review is the pharmacological diversity of the included interventions. Bumetanide, arbaclofen, and valproate are grouped here under the shared mechanistic hypothesis of GABAergic modulation, but they act through substantially different pathways: bumetanide restores the inhibitory action of GABA via NKCC1 inhibition, arbaclofen is a direct GABA-B receptor agonist that enhances inhibitory signalling, and valproate is a broad-spectrum agent whose relationship to GABAergic transmission is more indirect and accompanied by numerous non-GABAergic mechanisms. Because bumetanide trials contribute the majority of the evidence base in this review (8 of 13 studies), our pooled, class-level estimates should be interpreted as most heavily influenced by bumetanide’s specific effect profile, diluted or modified by the smaller and mechanistically distinct arbaclofen and valproate literatures, rather than as evidence of a shared “class effect.” This is consistent with the discrepancy between domains: the domains showing nominal signals, global severity and adaptive behaviour, are also the domains in which bumetanide trials predominate, whereas domains with more balanced contributions across agents (e.g., social communication, to which both arbaclofen trials and several bumetanide trials contributed) showed no signal at all. Formal drug-stratified analyses are needed before any claim about the efficacy of “GABA modulation” as a unified therapeutic strategy can be made with confidence.

This mechanistic heterogeneity may also help explain the uniformly null result for irritability (Hedges’ g = −0.07, 95% CI −0.39 to 0.24). As discussed above, GABAergic modulation is not expected to act unidirectionally on affective and behavioural symptoms: some patients may show reduced irritability with enhanced inhibitory signalling, while others may show no change or paradoxical worsening. A pooled average across such divergent individual responses, and across mechanistically distinct drugs, would be expected to approximate the null result observed here even if genuine, opposite-direction effects exist within subgroups of patients. This possibility cannot be tested with the aggregate data available in the present review but is a specific, testable hypothesis for future individual-patient-data analyses.

### 4.3. Risk of bias is the dominant limitation on certainty across domains

Methodological quality was variable across the thirteen included trials: only two were judged at overall low risk of bias, four had some concerns, and seven were judged at high risk, meaning approximately 85% of the evidence base carried some concern or high risk of bias overall. Bias in selection of the reported result (Domain 5) and in outcome measurement (Domain 4) were the principal drivers of these judgments. This pattern is directly relevant to interpreting the two nominal positive findings. Where studies assessed similar outcomes using multiple instruments, selective reporting of favourable results could have contributed to an apparent positive effect. Although our prespecified hierarchical approach to instrument selection reduced this risk (Supplementary Tables S4–S5), it could not eliminate bias already present in the original trial reports. GRADE certainty was correspondingly moderate at best for any outcome (global autism severity and adaptive behaviour), low for functional communication, RRBs, and irritability, and very low for social communication.

The restricted and repetitive behaviours (RRB) domain illustrates a related, evidence-availability limitation rather than a risk-of-bias limitation: Hollander et al. (2005) reported a large, statistically significant improvement in repetitive behaviours with divalproex (p = .037; Cohen’s d = 1.616), but could not be incorporated into the pooled estimate because C-YBOCS outcome data were presented graphically without extractable numerical values. Given the substantial magnitude of this excluded effect, even a very small sample, the RRB pooled estimate presented here (Hedges’ g = −0.21, 95% CI −0.51 to 0.09) should be regarded as a conservative, likely-attenuated estimate of the true effect in this domain.

### 4.4 Comparison with existing literature and clinical guidance

Our null findings for social communication and functional communication are consistent with current clinical guidance, which does not support pharmacological therapy for the core social-communication or communication deficits of ASD (Howes et al., 2017), and with the mixed individual-trial literature. For example, Veenstra-VanderWeele et al., 2016, a phase II trial of arbaclofen did not demonstrate improvement in its prespecified primary outcome of social withdrawal despite favourable findings on selected secondary and exploratory outcomes. Functional communication in particular is influenced by developmental maturation, intellectual ability, baseline language level, and concurrent behavioural or educational interventions, any of which could obscure a modest pharmacological effect within the short duration of the included trials (8 weeks to 6 months). Similarly, our null RRB and irritability findings are broadly consistent with guidance that continues to emphasize behavioural and developmental interventions for restricted and repetitive behaviours, and that reserves pharmacotherapy for these domains primarily for associated symptoms such as aggression, where agents with different mechanisms, such as risperidone and aripiprazole, have established efficacy (Howes et al., 2017).

### 4.5 Strengths of the Study

This review has several strengths. First, it focused specifically on randomized controlled trials, thereby limiting the influence of uncontrolled confounding and regression to the mean. Second, the analysis attempted to categorize heterogeneous scales into clinically relevant domains to facilitate pooled analysis. ASD trials frequently use different instruments to assess overlapping constructs; pooling conceptually similar measures using standardized mean differences therefore allowed studies using different scales to contribute to the same domain. The use of a hierarchical framework when multiple measures from the same study were available also reduced the risk of double counting and preserved the independence of observations. Critically, we applied and transparently reported a multiplicity correction across the six prespecified domains, that materially changes the interpretation of this evidence base relative to what an uncorrected analysis alone would suggest.

### 4.6 Limitations

Several limitations should be considered when interpreting these findings. The number of studies contributing to individual outcome domains was generally small, ranging from four studies for adaptive behaviour to nine study estimates for global autism severity, resulting in greater uncertainty for some pooled estimates, particularly those for social communication and restricted and repetitive behaviours. The limited number of studies also reduced the power to detect drug-specific effects, between-study heterogeneity, and reliable assessment of publication bias. Although all included interventions were classified as GABA-modulating therapies, they differ substantially in their pharmacological mechanisms; therefore, the pooled estimates should be interpreted as reflecting the overall effects of GABA-modulating interventions rather than the efficacy of any individual agent. In addition, ASD is a clinically heterogeneous disorder, and differences in age, developmental stage, intellectual ability, language ability, symptom severity, co-occurring conditions, and concomitant therapies may influence treatment response. The available evidence did not permit meaningful subgroup analyses, and the pooled estimates therefore represent average effects across diverse study populations. Outcome measures also varied across studies, necessitating the use of standardized mean differences to pool conceptually similar constructs. Although this approach facilitated quantitative synthesis, standardized mean differences are inherently less clinically interpretable than the original scales. Finally, selective outcome reporting and publication bias cannot be excluded. Funnel plots were not considered reliable for several outcomes because of the limited number of studies, and unpublished negative or null trials may therefore have influenced the pooled estimates.

### 4.7 Implications for Future Research

The findings support continued investigation of GABA-modulating pharmacotherapies in ASD while highlighting several priorities for future research. First, adequately powered randomized controlled trials are needed to improve the precision of treatment effect estimates, particularly for domains such as social communication and restricted and repetitive behaviours where current evidence remains inconclusive. Second, greater consistency in outcome measurement is needed. The use of validated, standardized instruments with clearly defined primary and secondary endpoints would improve comparability across studies and facilitate future evidence synthesis. As additional randomized trials become available, drug-specific analyses may help determine whether individual GABA-modulating agents demonstrate differential efficacy across ASD symptom domains. Finally, future studies incorporating biological or clinical markers may help identify patient subgroups most likely to benefit from specific GABA-modulating interventions and further clarify the role of GABA-related mechanisms in ASD (Cellot and Cherubini, 2014; Rubenstein and Merzenich, 2003).

## Conclusion

In this systematic review and meta-analysis of thirteen randomized controlled trials, GABA-modulating pharmacotherapies (bumetanide, arbaclofen, and valproate) did not demonstrate a robust treatment effect in any of six prespecified ASD symptom domains once correction for multiple comparisons was applied. Nominal, unadjusted improvements were observed for global autism severity and adaptive behaviour. These findings do not support a conclusion that GABA-modulating pharmacotherapies, considered as a class, produce broad or reliably domain-selective improvements in ASD. They are, however, consistent with a narrower and more defensible hypothesis: that any true therapeutic effect of GABAergic modulation in ASD is likely agent-specific and patient-specific rather than a shared “class effect,” given the substantial mechanistic differences among bumetanide, arbaclofen, and valproate and the biological plausibility of bidirectional, patient-dependent responses to GABAergic modulation. However, they suggest that the effects of GABA-modulating interventions are not uniform across ASD symptom domains and reinforce the importance of evaluating domain-specific outcomes rather than relying solely on global measures when assessing pharmacological interventions for ASD. The findings should be interpreted cautiously because of the limited number of studies contributing to several outcome domains, pharmacological and clinical heterogeneity, variability in outcome measures, and the small magnitude of the observed treatment effects. Additional adequately powered randomized controlled trials using standardized domain-specific outcome measures are needed to determine whether individual GABA-modulating interventions provide clinically meaningful benefits across specific ASD symptom domains.

## Data Availability

All data produced in the present work are contained in the manuscript

## Author Contributions

**Sanjana Palakodeti:**

Conceptualization, Methodology, Investigation, Data curation, Formal analysis, Visualization, Writing – original draft, Writing – review & editing, Project administration, Supervision.

**Kirti Kumar Hinduja:**

Conceptualization, Methodology, Investigation, Data curation, Visualization, Writing – original draft, Writing – review & editing, Project administration.

**Garima Mishra:**

Conceptualization, Investigation, Data curation, Visualization, Writing – original draft, Writing – review & editing.

**Subhiksha R:**

Investigation, Data curation, Visualization.

**Abhigna Pabbaraju:**

Investigation, Data curation.

**Bagath Srinivasan Balaji:**

Methodology, Investigation, Data curation, Visualization, Writing – original draft.

**Tejasvi Parmar:**

Data curation.

## Data Availability Statement

The data supporting the findings of this study are available within the article and its supplementary material. Additional extracted data are available from the corresponding author upon reasonable request.

## Ethics Approval

Not applicable

## Consent to participate

Not Applicable

## Consent for Publication

Not Applicable

