## Supplementary Material for "Domain-Specific Effects of GABA-Modulating Pharmacotherapies in Autism Spectrum Disorder: A Systematic Review and Meta-Analysis of Randomized Controlled Trials"

**Supplementary Table S1. Full Search Strategies**

| **Database** | **Search Strategy** |
| --- | --- |
| Pubmed | ("Autism Spectrum disorder" OR Autis* OR ASD OR Kanner OR Asperger OR "Asperger Syndrome" OR PDD OR "Pervasive developmental disorder" OR "Autistic Disorder"[Mesh]) AND (baclofen OR arbaclofen OR bumetanide OR riluzole OR "GABA Agents"[Mesh] OR "GABA agonist*" OR "GABA antagonist*" OR "GABA modulator*")<br><br>("Autism Spectrum disorder" OR Autis* OR ASD OR Kanner OR Asperger OR "Asperger Syndrome" OR PDD OR "Pervasive developmental disorder" OR "Autistic Disorder"[Mesh]) AND (“valproate” OR “valproic acid”). Filters - 2024 to 2026 |
| Cochrane | #1: ("Autism Spectrum disorder" OR ASD OR Kanner OR Asperger OR "Asperger Syndrome" OR PDD OR "Pervasive developmental disorder")<br>#2: MeSH Descriptor: [Autistic Disorder] explode all trees<br>#3: (baclofen OR arbaclofen OR bumetanide OR riluzole OR "GABA agonists" OR "GABA antagonists" OR "GABA modulators" OR "acamprosate")<br>#4: MeSH Descriptor: [GABA Modulators] explode all trees<br>Final Search: (#1 OR #2) AND (#3 OR #4) |

| | |
|---|---|
| Scopus | ( "Autism Spectrum Disorder" OR Autis* OR ASD OR Kanner OR Asperger OR "Asperger Syndrome" OR PDD OR "Pervasive Development Disorder" OR "Autistic Disorder" ) AND ( baclofen OR arbaclofen OR bumetanide OR riluzole OR "GABA agents" OR "GABA agonist*" OR "GABA antagonist*" OR "GABA modulator*" )<br>Filters: Relevant keyword filters applied |
| Embase | ( "Autism Spectrum Disorder" OR Autis* OR ASD OR Kanner OR Asperger OR "Asperger Syndrome" OR PDD OR "Pervasive Development Disorder" OR "Autistic Disorder" ) AND ( baclofen OR arbaclofen OR bumetanide OR riluzole OR "GABA agents" OR "GABA agonist*" OR "GABA antagonist*" OR "GABA modulator*" )<br>Filters: English language |

**Supplementary Table S2. Classification of Outcome Measures**

| Outcomes | Domain | Instruments |
|---|---|---|
| **Primary Outcomes** | 1. Global autism severity | CARS, CARS-2, SRS, SRS-2 |
| | 2. Social functioning / social communication | SRS-SCI, VABS Socialization, ABC-SW, ABC Interaction, SSRS, ADOS-B |
| | 3. Functional communication | VABS Communication, ADOS-A, ABC-L |
| | 4. Restricted and repetitive behaviours | RBS-R, SRS RRB, ABC Stereotypies, C-YBOCS, ADOS-RRBs |
| | 5. Adaptive functioning | VABS Daily Living/Composite, ABC-SSH |
| **Secondary Outcomes** | Irritability | ABC-I |

**Supplementary Table S3. Rationale for Outcome-Domain Classification and Scale Combination**

| Outcome domain | Instruments included | Construct represented by the instruments | Rationale for combining scales within the domain |
|---|---|---|---|
| **Global autism severity** | Childhood Autism Rating Scale (CARS/CARS-2); Social Responsiveness Scale (SRS/SRS-2) | Overall severity of autistic symptoms and global autism-related impairment | These instruments provide global or broad measures of autism symptom severity rather than assessing a single isolated symptom domain. Although the scales differ in item content and scoring range, their common purpose is to quantify overall autism-related symptom burden. Standardized mean differences (Hedges' *g*) were therefore used to permit pooling across different measurement scales. When multiple eligible measures were available within the same study, the CARS was prioritized over the SRS because it was considered more directly focused on overall autism symptom severity. |

| | | | |
|---|---|---|---|
| **Social communication / social functioning** | SRS Social Communication and Interaction (SRS-SCI); Vineland Adaptive Behavior Scales (VABS) Socialization; Autism Diagnostic Observation Schedule (ADOS) social-communication component; Aberrant Behavior Checklist (ABC) Social Withdrawal/Relating; Social Skills Rating System (SSRS) | Social communication, social interaction, social responsiveness, and social functioning | The instruments capture overlapping aspects of social functioning and social-communication impairment, including reciprocal social interaction, social responsiveness, and communication-related social behaviour. Because the measures use different scales and scoring systems, SMDs were used to standardize treatment effects. The SRS-SCI was prioritized when available because it specifically assesses social communication and interaction deficits corresponding to the core social domain of ASD. |
| **Functional communication** | VABS Communication; ADOS Communication component; ABC Language | Functional language and communication abilities | These measures assess communication-related functioning, including expressive/receptive communication or observable communication behaviours. Although the instruments differ in format and assessment method, they address the common construct of communication functioning. SMDs were therefore used to combine effects across instruments. Measures specifically assessing communication were prioritized over broader global or behavioural measures. |

| | | | |
|---|---|---|---|
| **Restricted and repetitive behaviours (RRBs)** | Repetitive Behavior Scale–Revised (RBS-R); SRS Restricted Interests and Repetitive Behavior (RRB) subscale; ABC Stereotypy; Children's Yale-Brown Obsessive Compulsive Scale (C-YBOCS) compulsion subscale; ADOS RRB-related component | Restricted interests, repetitive behaviours, stereotyped behaviours, compulsive/repetitive behaviours | These instruments assess overlapping manifestations of restricted, repetitive, stereotyped, or compulsive behaviour, representing the RRB domain of ASD. SMDs permit pooling despite differences in scale structure and scoring. Selection prioritized measures that were most directly specific to RRBs and autism-related repetitive behaviour. For example, the RBS-R was preferred over broader measures when both assessed the same construct. |
| **Adaptive functioning / adaptive behaviour** | VABS Daily Living Skills/Adaptive Behavior Composite; ABC Self-Help/Self-Sufficiency | Practical, daily-living, self-care, and adaptive functioning | These measures assess functional abilities required for everyday independent functioning, including self-care and daily living skills. Although the instruments differ in structure, they represent the broader construct of adaptive functioning. SMDs were therefore used to standardize effects across instruments. Measures specifically assessing adaptive functioning were prioritized over measures of core ASD symptom severity. |
| **Irritability** *(secondary outcome)* | ABC Irritability (ABC-I) | Irritability, aggression, tantrums, and related behavioural dysregulation | The ABC-I was used because it is a standardized and commonly used measure of irritability in ASD pharmacological trials. As the same instrument was used across contributing studies, the outcome was conceptually and methodologically consistent across trials. |

**General principle for scale combination:** Different instruments were pooled within each prespecified domain when they were judged to assess the same overarching clinical construct. Because instruments differed in measurement scales, scoring ranges, and units, treatment effects were expressed as standardized mean differences using Hedges' *g*. To avoid double counting participants, only one outcome measure per domain per study was selected for the primary meta-analysis when multiple eligible measures were reported. Selection followed a prespecified hierarchy: (1) the most construct-specific instrument; (2) autism-specific measures over general behavioural or adaptive measures; (3) symptom-severity measures over diagnostic algorithm subscales; (4) clinician-rated measures over caregiver-rated measures when construct coverage was comparable; and (5) the most frequently used instrument across studies when otherwise equivalent. The direction of effect was harmonized before pooling so that negative effect sizes consistently favoured the intervention.

**Supplementary Table S4. Prespecified Outcome Selection Hierarchy**

Several included studies reported multiple instruments assessing similar constructs within the same outcome domain. To maintain statistical independence by including only one effect estimate per study for each meta-analysis, a prespecified hierarchical framework was applied to select the most appropriate outcome measure. This was the hierarchy followed:

1. Most construct-specific measure.
2. Autism-specific scales over general behavioural/adaptive scales.
3. Symptom severity scales over diagnostic algorithm subscales.
4. Clinician-rated over caregiver-rated measures when comparable.
5. Most frequently used measure when multiple equivalent options remain.

**Supplementary Table S5. Study-specific Scale Selection Decisions**

The outcome measure selected for each study when multiple eligible instruments were available within the same outcome domain, together with the rationale for each decision. These selections were made according to the prespecified hierarchy described in Table S4.

| **Domain** | **Author** | **Available Scales** | **Chosen** | **Reason** |
|---|---|---|---|---|
| **RRBs** | Parellada | ABC-Stereotypy, SRS2P RRB | SRS-RRB | More autism-specific than ABC-Stereotypy |
| | Vanderweele | ABC- Stereotypy, Vineland Maladaptive Behaviors Subscale | ABC-Stereotypy | More specific than VABS Maladaptive |
| | Dai | RBS-R, ADOS R | RBS-R | More specific than ADOS R |

| | | | | |
|---|---|---|---|---|
| **Social Communication** | Parellada | VABS socialization,<br>SRS-P SCI,<br>ABC-social withdrawal | SRS-P SCI | The SRS-SCI score was specifically designed to measure DSM-5 Criterion A (social communication and interaction deficits) and maps directly onto the modern conceptualization of ASD social symptoms. In contrast, VABS Socialization assesses adaptive social functioning and daily-life social skills.<br><br>ABC Social Withdrawal/Relating measures social disengagement but was not developed to operationalize DSM-5 social communication deficits |
| **Global Function** | Fuentes | CARS2, SRS2 | CARS | CARS>SRS |
| | Lemonnier 2017 | CARS, SRS | CARS | CARS>SRS |
| | Dai | CARS, SRS | CARS | CARS>SRS |

**Supplementary Table S6. Holm–Bonferroni Step Down adjustment for multiple comparisons across prespecified outcome domains**

| **Outcome** | **Hedges' g** | **95% CI** | **Unadjusted p** | **Holm-adjusted p** | **Significant after Holm adjustment?** |
|---|---|---|---|---|---|
| Adaptive behaviour | −0.09 | −0.15 to −0.02 | .023 | **.138** | No |
| Global autism severity | −0.25 | −0.46 to −0.03 | .028 | **.140** | No |
| Social communication | −0.26 | −0.55 to 0.04 | .077 | .308 | No |
| RRBs | −0.21 | −0.51 to 0.09 | .126 | .378 | No |
| Irritability | −0.07 | −0.39 to 0.24 | .543 | 1.000 | No |
| Functional communication | −0.01 | −0.17 to 0.15 | .869 | 1.000 | No |

CI, confidence interval; g, Hedges' g. Holm-adjusted p-values were calculated across the six prespecified outcome-level hypothesis tests using the Holm–Bonferroni step-down procedure. P-values were ordered from smallest to largest before adjustment. The Holm multiplier was calculated as $(m - i + 1)$, where $m = 6$ outcome domains and i denotes the rank of the ordered p-value. Adjusted p-values were constrained to a maximum of 1.00. Statistical significance was defined as $p < .05$. The multiplicity adjustment was applied to p-values only and did not modify the pooled effect estimates or their 95% confidence intervals.

### Supplementary Figures

### Figure S1. Sensitivity Analysis of Global Autism Severity Domain Excluding Zhang 2020

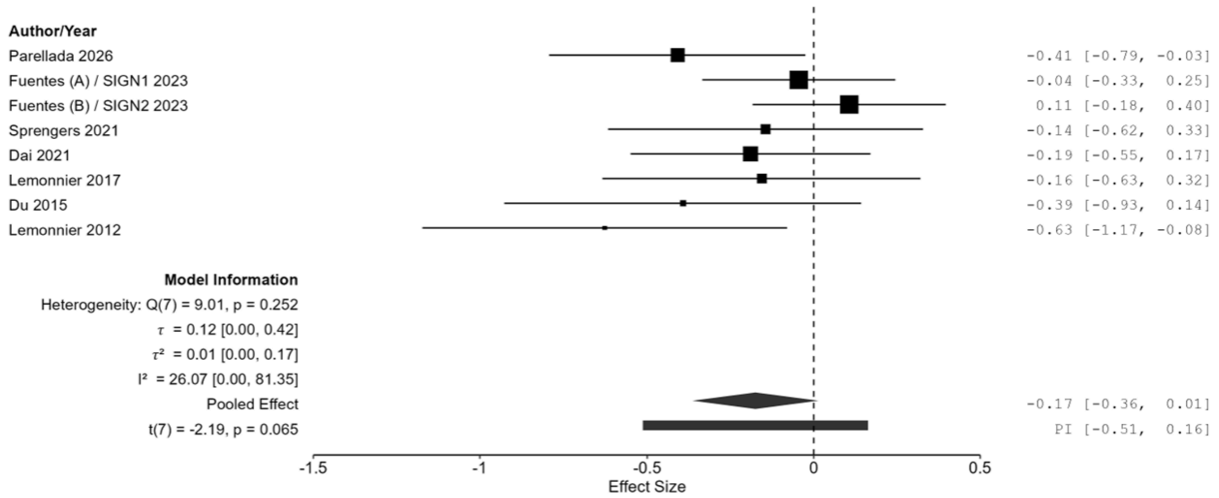

**Figure S2. Funnel plot for Group 1 – Global Autism Severity**

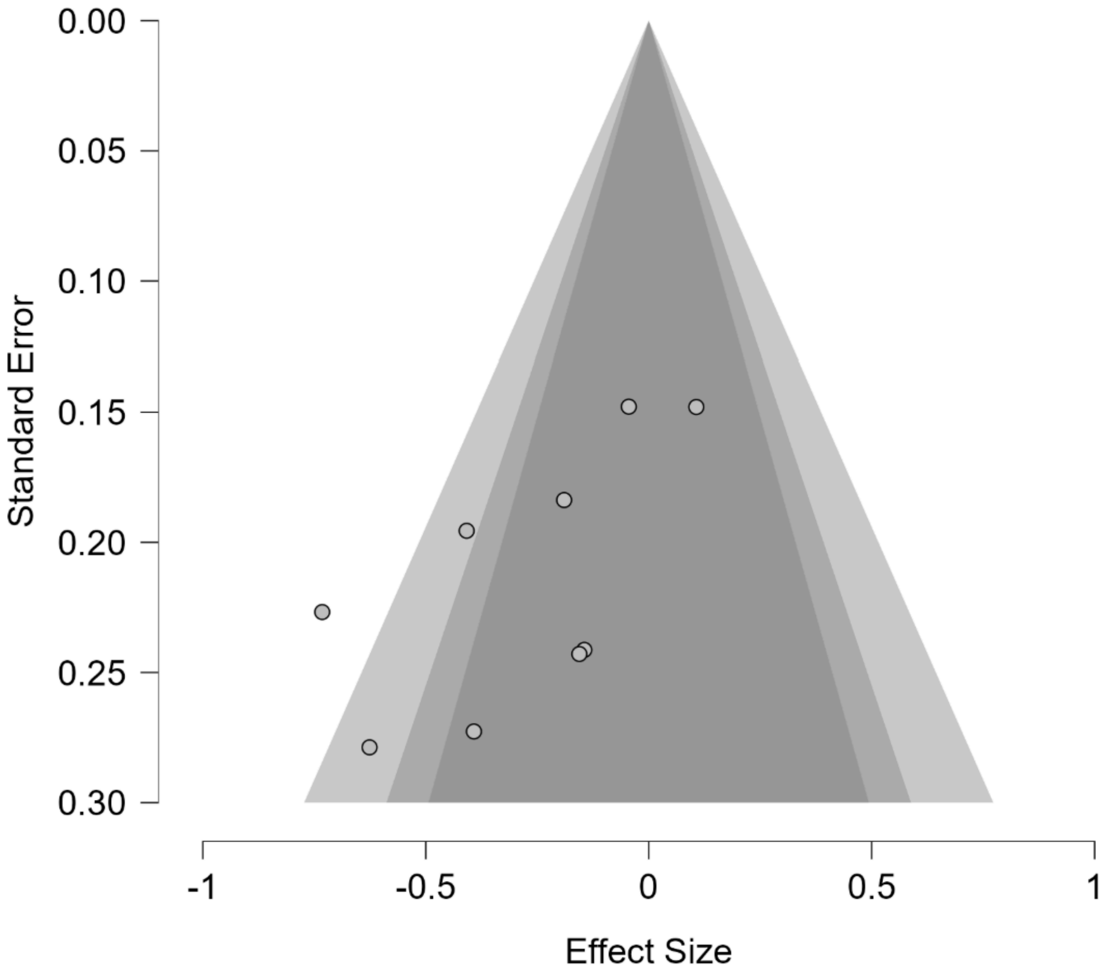
